# A detectability floor for treatment-response labels in transcriptomics: a methodological audit using rheumatoid arthritis as a test case

**DOI:** 10.64898/2026.10.04.26364671

**Authors:** Haitao Yang, Yan Huang, Jianghua Xu, Suo Zhang, Rui Liao

## Abstract

**Background:** Treatment-response labels in rheumatoid arthritis (RA) transcriptomics are used to stratify patients, yet before-and-after studies rarely yield a reproducible treatment signal. We audited whether such labels are detectable within the designs that define them, using disease-state labels as an internal comparator.

**Methods:** Eight public paired datasets (juvenile dermatomyositis n=10; RA 28 pairs; lupus nephritis n=10; controlled exposures; a single-cell dose series) were analysed under a pre-registered verdict taxonomy. Each label’s effect size (Cohen’s dz, 95% CI) was compared with the minimum detectable effect (MDE) of its own design; response labels in current use were audited in every analysable cohort.

**Results:** Disease-state labels were detectable: interferon fell in 10/10 juvenile dermatomyositis patients (p=0.002). No exposure label cleared its floor; a drug-matched methotrexate prior was nominally significant yet below it (dz=-0.80; 95% CI −1.51 to −0.09; floor 1.0). Of 15 audited cohorts, 1 was detected as published, 3 bounded below their floor and 11 unresolved at it; 3 pointed opposite to the claim, each below its own floor, none reversed. The largest cohort gave dz=-0.45 (95% CI −0.82 to −0.07) and the pooled estimate +0.26 (95% CI −0.18 to 0.70; I²=69%), both below their floors.

**Conclusions:** Within these cohorts, exposure and response-prediction labels did not reach the detection floors of their designs, whereas disease-state labels did. Reporting both quantities would let a null be read as a bound; what a below-floor label implies for stratification or benchmarking is not settled here.

**Trial registration:** Not applicable.

## Introduction

Molecular stratification in autoimmune rheumatic disease has outpaced treatment-response prediction. Interferon programmes in SLE [1], TNF and inflammatory programmes in RA and axial spondyloarthritis, and fibrotic programmes in SSc are reproducible and increasingly used to describe patients; these labels describe what a cell or tissue is. Before-and-after transcriptomic studies—the design that asks what a treatment did to that same tissue—have been far less reproducible. In SSc, few treatment-associated genes survive correction across trial biopsies [2]; in SLE, conventional-dose glucocorticoids often fail to suppress interferon-stimulated genes over long follow-up [3]; in axial spondyloarthritis, bulk responses to TNF blockade are modest and responder-dependent [4]. Mechanism-specific biologics may differ: belimumab modulates molecular signatures in SLE [5]. Whether the pattern extends across drugs, tissues and platforms has not been systematically assessed.

A similar asymmetry is evident in the virtual-cell literature. Transcriptomic foundation models such as scGPT [6] and Geneformer [7] transfer representations across donors, tissues and protocols [6–8], and perturbation-scale models transfer to patient-derived organoids and biopsies [9]. These successes share a structural feature: the labels encode large, controlled displacements—cell identity, knockout aftermath, high-dose drug exposure. The clinically most consequential application asks something different: given a patient’s cells before therapy, forecast what a conventional treatment will do to them. Performance there has been unstable: deep-learning models do not consistently outperform simple linear baselines when perturbation effects must be predicted [10]. One reading is architectural; another, untested in the labels themselves, is that the label differs in kind—a modulation within a state under clinical rather than laboratory control. On that reading the failure is the label’s rather than the model’s, checkable before modelling.

Labels differ in the magnitude of the biological displacement they encode: at one end, state identity; in the middle, strong controlled perturbation; at the other, clinical conventional exposure, where the drug is only one determinant of the measured state alongside dose, timing, concomitant therapy, adherence, tissue and baseline activity. We call this the effect-size spectrum of supervision labels, and hypothesise that a label’s detectability in a given design declines along it. The spectrum is a property of the label, detectability the product of label and design; movement along it is necessary but not sufficient, since sampling interval and prior– context match also move a label independently of effect size.

We hypothesised that within the same patients, tissues and scoring framework, disease-state labels would be detectable, whereas treatment-exposure labels would not reach the floors of their own designs. Detectability is treated as a joint function of prior–context match, exposure intensity, tissue purity and sampling design, so restoring any one should render a label detectable, whereas escalating dose alone in a mismatched context should not. Using public paired transcriptomics (Table 1), we (i) contrast disease-state and exposure labels in the same patients in juvenile dermatomyositis [11]; (ii) show that exposure-disease correlations in mixed designs are composition and scoring artefacts [12–14]; (iii) replicate the exposure nulls in RA [15] and lupus nephritis [16], cross-checked against the authors’ own analyses and a drug-matched prior; (iv) bound detectability from below with controlled exposures [17–19] and a single-cell dose series [20,21]; and (v) audit labels in current use, one benchmark trajectory and a cross-disease panel [22–28]. Effect sizes are reported against each design’s minimum detectable effect, so negative findings are bounds, not absences.

**Table 1.**
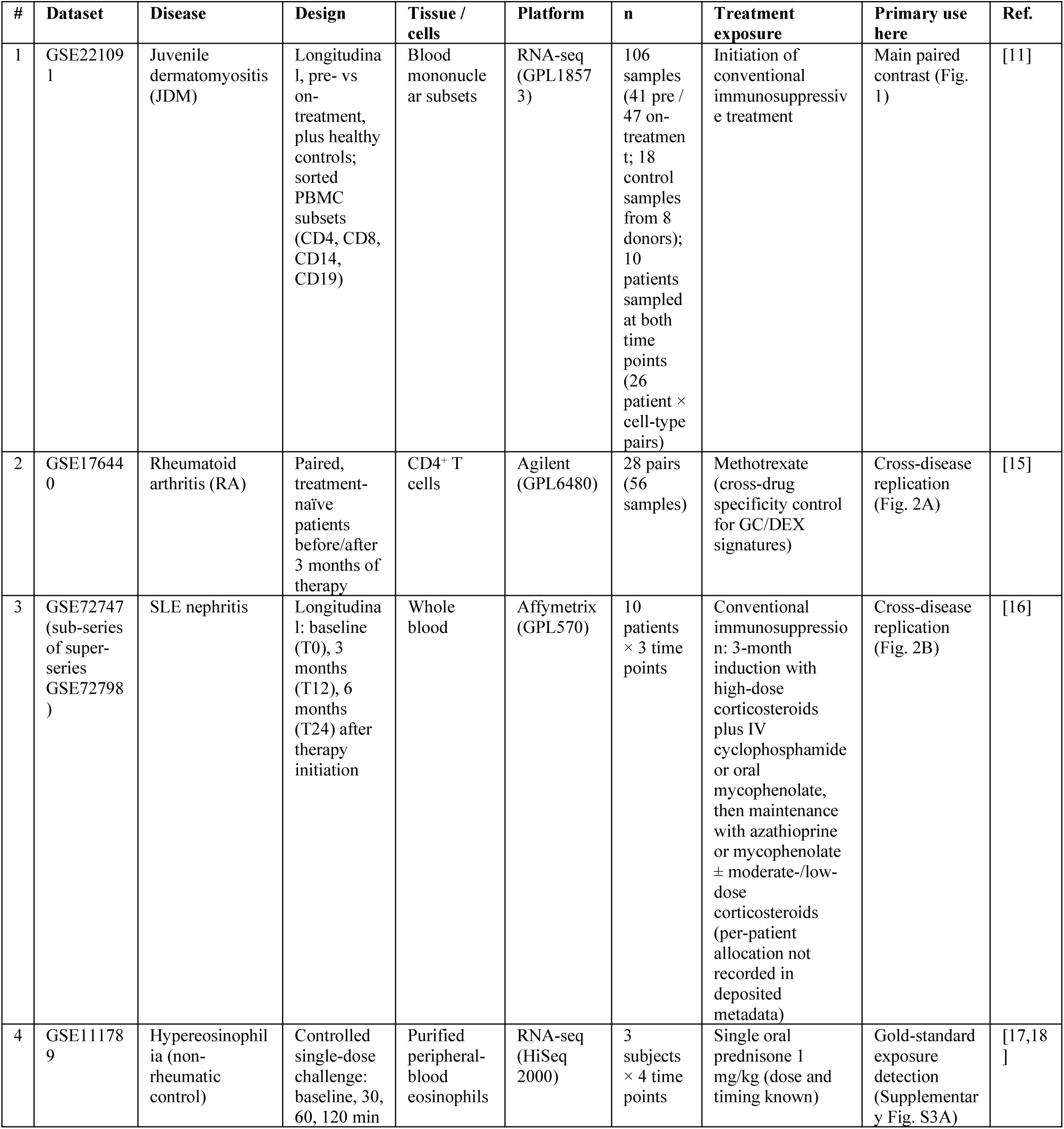
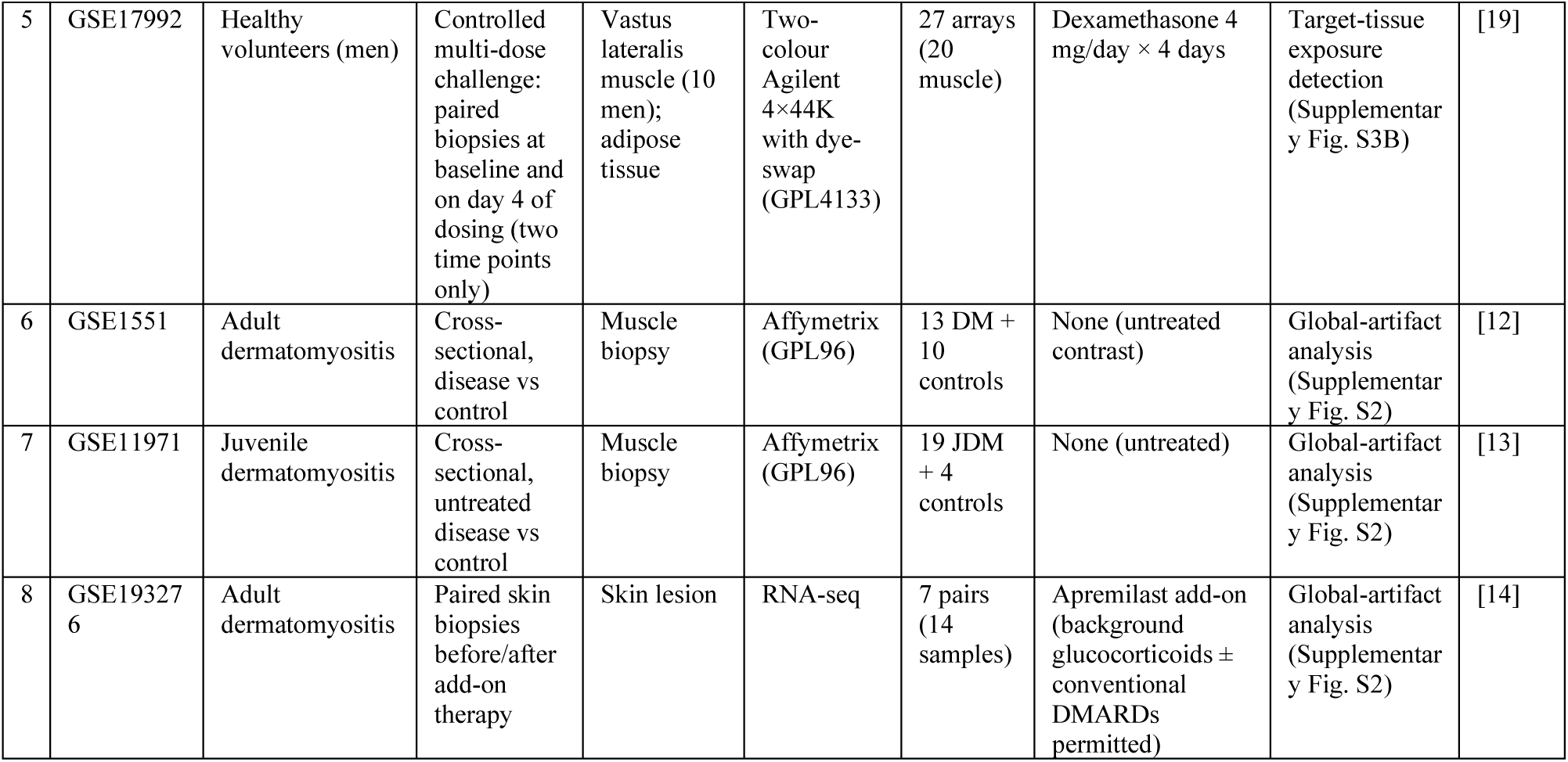
Datasets, designs and exposures.

| **#** | **Dataset** | **Disease** | **Design** | **Tissue / cells** | **Platform** | **n** | **Treatment exposure** | **Primary use here** | **Ref.** |
|---|---|---|---|---|---|---|---|---|---|
| 1 | GSE221091 | Juvenile dermatomyositis (JDM) | Longitudinal, pre- vs on-treatment, plus healthy controls; sorted PBMC subsets (CD4, CD8, CD14, CD19) | Blood mononuclear subsets | RNA-seq (GPL18573) | 106 samples (41 pre / 47 on-treatment; 18 control samples from 8 donors); 10 patients sampled at both time points (26 patient × cell-type pairs) | Initiation of conventional immunosuppressive treatment | Main paired contrast (Fig. 1) | [11] |
| 2 | GSE176440 | Rheumatoid arthritis (RA) | Paired, treatment-naïve patients before/after 3 months of therapy | CD4<sup>+</sup> T cells | Agilent (GPL6480) | 28 pairs (56 samples) | Methotrexate (cross-drug specificity control for GC/DEX signatures) | Cross-disease replication (Fig. 2A) | [15] |
| 3 | GSE72747 (sub-series of super-series GSE72798) | SLE nephritis | Longitudinal: baseline (T0), 3 months (T12), 6 months (T24) after therapy initiation | Whole blood | Affymetrix (GPL570) | 10 patients × 3 time points | Conventional immunosuppression: 3-month induction with high-dose corticosteroids plus IV cyclophosphamide or oral mycophenolate, then maintenance with azathioprine or mycophenolate ± moderate-/low-dose corticosteroids (per-patient allocation not recorded in deposited metadata) | Cross-disease replication (Fig. 2B) | [16] |
| 4 | GSE111789 | Hypereosinophilia (non-rheumatic control) | Controlled single-dose challenge: baseline, 30, 60, 120 min | Purified peripheral-blood eosinophils | RNA-seq (HiSeq 2000) | 3 subjects × 4 time points | Single oral prednisone 1 mg/kg (dose and timing known) | Gold-standard exposure detection (Supplementary Fig. S3A) | [17,18] |
| 5 | GSE17992 | Healthy volunteers (men) | Controlled multi-dose challenge: paired biopsies at baseline and on day 4 of dosing (two time points only) | Vastus lateralis muscle (10 men); adipose tissue | Two-colour Agilent 4×44K with dye-swap (GPL4133) | 27 arrays (20 muscle) | Dexamethasone 4 mg/day × 4 days | Target-tissue exposure detection (Supplementary Fig. S3B) | [19] |
| 6 | GSE1551 | Adult dermatomyositis | Cross-sectional, disease vs control | Muscle biopsy | Affymetrix (GPL96) | 13 DM + 10 controls | None (untreated contrast) | Global-artifact analysis (Supplementary Fig. S2) | [12] |
| 7 | GSE11971 | Juvenile dermatomyositis | Cross-sectional, untreated disease vs control | Muscle biopsy | Affymetrix (GPL96) | 19 JDM + 4 controls | None (untreated) | Global-artifact analysis (Supplementary Fig. S2) | [13] |
| 8 | GSE193276 | Adult dermatomyositis | Paired skin biopsies before/after add-on therapy | Skin lesion | RNA-seq | 7 pairs (14 samples) | Apremilast add-on (background glucocorticoids ± conventional DMARDs permitted) | Global-artifact analysis (Supplementary Fig. S2) | [14] |

Two appraisals of the rheumatoid arthritis treatment-response literature bear directly on this question: a critical appraisal of the machine-learning models built for that purpose found most of them at unclear or high risk of bias against PROBAST and TRIPOD [29], and a systematic review of the clinical prediction models for methotrexate outcomes found that none met the minimum sample-size requirement for model development [30]. Both address the model— whether it was developed on enough events to be stable and reported adequately; neither addresses the label the model scores. A label deposited with a published result is a claim about an effect size, carrying a detectability condition set by the size of the cohort that measured it, not by the adequacy of any model fitted to it. Our unit of analysis is therefore the paper and its headline label, compared against the floor of the cohort that produced it; where an existing appraisal reports an underpowered model, we ask the complementary question of whether the signal the model was fitted to was measurable at that cohort’s size—the two can disagree, because a label can carry a real effect that the cohort was never able to detect.

No model is evaluated here. The study asks whether the labels that treatment-response evaluations rest on are measurable in the designs defining them. If the hypothesis holds, part of the negative before-and-after literature would reflect a design floor, and design modification rather than cohort expansion would be the implication to be tested prospectively.

## Methods

### Study design and datasets

Eight public transcriptomic datasets were assembled to score disease-state and exposure labels within the same patients, tissues and scoring framework, to replicate nulls across diseases and bound detectability from below with two controlled experiments (Table 1; Table S1). GSE72747 is the lupus-nephritis immunosuppression arm of super-series GSE72798, whose linked publication reports the IFNα-kinoid arm [16]; its regimen is documented at protocol level, per-patient drug allocation is not.

### Gene signatures and prior gene sets

Three label classes are distinguished by intent rather than biology. **Disease-state labels** encode a tissue’s position on its pathological axis; **exposure fingerprints** encode the drug-specific component of post-treatment change; **response-prediction labels** are measured at baseline and predict a future change quantity. Disease-state labels were represented by the union of the MSigDB Hallmark interferon-alpha and interferon-gamma response sets (224 genes) [31], with a TNF/inflammation signature as disease-activity control [31]. Exposure fingerprints were represented by four glucocorticoid-responsive sets of descending curation quality: a curated Gene Ontology set (GO:0071385, GO:0051384, 30 genes) [32]; a 10-gene canonical target set; a 1,149-gene dexamethasone-upregulated GEO set [33]; and an 887-gene NR3C1 ChIP-seq target set [33]. Two drug-specific checks complemented these: a methotrexate-specific prior (355 genes from GSE23687, 299 mapped) [34], deliberately not cell-type-matched; and a search of ten perturbation resources finding no apremilast signature. Symbols were mapped with each platform’s annotation table (rates in Supplementary Table S1).

### Signature scoring

Rank-based scoring was designated primary before any result was inspected, because rank scores are invariant to monotone per-gene transformations and do not track a sample’s global expression level. Two conventions were used by dataset family: a dataset-level rank score for GSE221091 and GSE111789, and the single-sample singscore [35] for GSE176440 and GSE72747. Sensitivity scorers (z-score-mean, an AUCell-type method, a cell-type-restricted rank variant) were computed alongside (Fig. S1).

### Statistical analysis

Paired within-patient contrasts used the two-sided Wilcoxon signed-rank test; two-group contrasts, the Mann-Whitney U test (equivalently reported as AUC). Effects are reported as Cohen’s dz with 95% CIs and, for GSE221091, as the variance explained (eta2). The primary GSE221091 analysis is at patient level (n=10 pairs); a patient × cell-type analysis (26 pairs) is sensitivity and overstates precision. Every exposure prior tested is disclosed with its result. For every design we computed the minimum detectable effect (MDE; smallest dz detectable at 80% power, two-sided alpha=0.05, exact non-central t) and classified each label as detected (|dz| at or above MDE), bounded below the floor (CI within ±MDE) or inconclusive (CI spanning it). The floor criterion is algebraically equivalent to |t| ≈ 2.80, both being functions of n and the pooled SD (Supplementary Methods); it is stated in effect-size units because a performance claim requires the magnitude a label carries. Two families were Benjamini-Hochberg corrected: the 12 empirical p values of the gold-standard experiment and the three subject-level specificity contrasts of GSE17992; clinical-cohort conclusions rest on effect-size bounds and CI placement, not nominal significance. The pipeline was calibrated by score-level injection (400 simulations) and paired dz values multiplied by J(n-1) as a small-sample check, with no verdict changing (Table S6).

### Empirical null for the gold-standard experiment

For GSE111789 each signature’s paired change was assessed against an expression-matched null: 300 random gene sets matched on size and expression decile, the empirical p value being the fraction whose absolute mean change met or exceeded the observed.

### Positive-control dose-response analysis (sci-Plex 2)

As a positive control we analysed the sci-Plex 2 dexamethasone dose series in A549 cells (0.1-100 microM, 24 h; 8,064 cells; 914 vehicle), from scPerturb [21] and outside the eight study datasets. Per-cell counts were normalised (CP10K, log1p) and scored with rank-based singscore [35] over a 9,516-gene universe, each dose against vehicle wells. Each dose carries six wells; the primary read-out is pseudobulk at well level (dose 100 versus vehicle, Welch t-test; exact two-sample MDE at n=6), with cell-level contrasts as sensitivity. For the interval axis (Fig. 3B) these results were combined with archived subject-level results for GSE111789 and GSE17992; the paradigms differ in unit and estimand and no extrapolation is implied.

### Label audits in current use

The ImmunoSTATE benchmark label [36] — a 14-gene CXCL13-positive T-cell module in synovial-fluid single-cell data from nine RA patients before and after tofacitinib or adalimumab (GSE296117 [37], 100,387 cells) — was re-derived as defined (4,907 CXCL13-positive T cells) and assessed at cell, patient and design-floor layers.

For the dominant conventional class, response signatures packaged with machine-learning predictors, candidates came from a documented two-database search with citation chasing (384 records, 132 after a transcriptome-by-response screen, 45 auditable), adjudicated line by line. Cohort-level audit covered 15 cohorts (rheumatoid arthritis primary, inflammatory bowel disease comparator), counted once per cohort; papers were audited in every analysable source cohort as deposited; five of the first paper’s eight cohorts do not permit exact reconstruction of the published subsets, a provenance gap flagged in Supplementary Table S1 rather than scored as F3. Per cohort the set score was compared between responders and non-responders (dz with percentile bootstrap CIs) against that cohort’s own MDE. Verdicts follow a pre-registered taxonomy: F1a (bounded below the floor), F1b (inconclusive, CI spanning it), F2 (detected sign-reversed relative to the published claim), F3 (not reproducible from the deposited record; excluded). F1a and F1b are reported separately, an interval spanning the floor being unresolved rather than null (Supplementary Methods 1.4). Each label is scored on the contrast it is deployed for; a diagnostic signature repurposed to a response task carries no direction expectation and is reported without a verdict. Paper-level summaries use random-effects (DerSimonian-Laird) combination with Q and I²; the same rules were applied to a cross-disease panel (Supplementary Methods, Tables S1-S3).

The search and adjudication were specified before any verdict was computed; the screening funnel is given in full in Supplementary Table S3. Fifteen cohorts from three source papers survived to assessment: eight from the largest audited paper [22], six from a second [25], and one inflammatory bowel disease comparator [26]. Further records were scored F3 (published subset not reconstructable from what was deposited) or reported without a verdict (a diagnostic-signature grouping, a single-cohort check and a test set), and are excluded from the auditable denominator rather than counted as failures. The taxonomy, including the requirement that a sign-reversal verdict presuppose detection, was registered before any verdict was computed.

### Mixed-sample artifact analysis

For GSE1551, GSE11971 and GSE193276, glucocorticoid and interferon scores were correlated under three scorers in two scopes (mixed samples; within patients); correlations were recomputed after regressing out each sample’s global expression level, and score variance decomposed into treatment status, cell type and patient identity (eta2).

### Patient and public involvement

Not applicable: this was a secondary analysis of public data, with no patient recruitment or contact.

## Results

### Disease-state labels are detectable; exposure labels are not, in the same samples (Fig. 1)

In GSE221091 [11], the interferon signature separated patients from controls and pre-from on-treatment samples (AUC 0.94; p=9.1 × 10-13). At patient level (n=10) the interferon score fell in 10/10 patients (dz=-3.0; p=0.002), treatment status explaining 59.5% of score variance versus 0.7-0.9% for glucocorticoid. Under the same scorer, the exposure label gave AUC 0.52 (p=0.77) and no consistent direction (dz=-0.22; p=0.56), although 9 of 11 on-treatment patients took oral glucocorticoids and all 11 methotrexate. A drug-matched methotrexate prior (drug-up in its source, so a drug signal would be positive) was nominally significant in the same patients (dz=-0.80; 95% CI −1.51 to −0.09) yet below its floor (1.0) and opposite to the drug-matched direction; its signal sat in the interferon-stimulated-gene component (dz=-2.85) and attenuated to −0.58 (CI −1.25 to 0.09) without it—consistent with disease-state information contributing to the prior rather than a methotrexate-specific fingerprint. This cohort has several favourable design features (treatment-naive tissue, induction-intensity treatment, sorted cells, complete pairing): the glucocorticoid label did not differ detectably between time points, and the nominally significant methotrexate signal remained below its floor.

**Figure 1.**
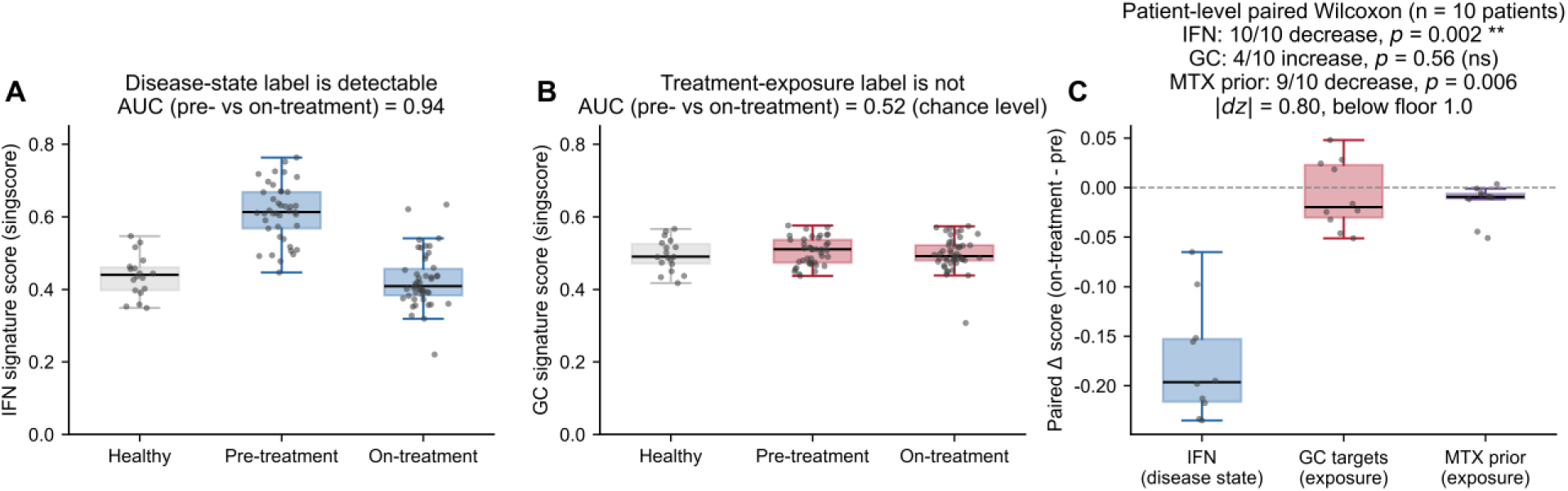
Disease-state and treatment-exposure labels in the same samples (GSE221091). (A) Interferon signature, pre-vs on-treatment (AUC 0.94). (B) Glucocorticoid-target signature, same contrast (AUC 0.52). (C) Patient-level paired changes: interferon fell in 10/10 patients (dz = −3.0; p = 0.002); the glucocorticoid score had no consistent direction (dz = −0.22; p = 0.56); the methotrexate prior fell in 9/10 (dz = −0.80; p = 0.006) yet did not clear its floor (1.0). Primary rank-based scorer; five-scorer agreement for interferon and glucocorticoid (Fig. S1). Alt text: Three panels. A, interferon separating pre-from on-treatment samples. B, glucocorticoid contrast, near-total overlap. C, paired changes: interferon falls in all ten; glucocorticoid flat; methotrexate prior below its floor.

### Mixed-sample correlations are composition artefacts (Supplementary Fig. S2)

In mixed designs, glucocorticoid and interferon scores correlated strongly (r up to 0.93) but collapsed within patients (r=-0.26), largely attributable to each sample’s global expression level.

### Exposure-label nulls replicate in RA and lupus nephritis (Fig. 2)

In 28 treatment-naive RA patients with paired CD4+ transcriptomes before and after three months of methotrexate, no signature changed reproducibly (p=0.41-0.96); a genome-wide paired re-analysis yielded 15 Benjamini-Hochberg-significant genes against 13 at q < 0.05 in the authors’ own table, none in any signature (enrichment p=0.11-0.73). A methotrexate-specific prior also fell below the floor (dz=0.015; 95% CI −0.36 to 0.39). In ten lupus nephritis patients sampled at baseline and 3 and 6 months after conventional immunosuppression, no exposure signature separated time points (p=0.32-0.43), and clinical responders showed no consistent interferon decline.

**Figure 2.**
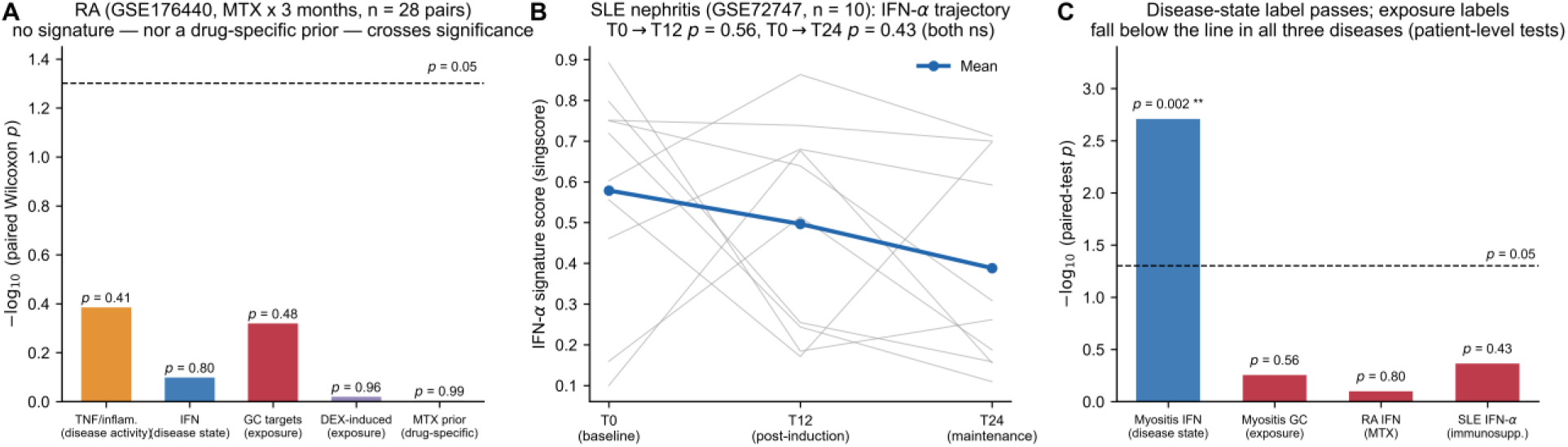
Exposure-label nulls across diseases. Paired changes with 95% CIs for four signatures in RA before and after methotrexate, and for three signatures in lupus nephritis across three time points, each against its design’s minimum detectable effect (dashed). No exposure label clears its floor; a methotrexate-specific prior is shown for comparison. Alt text: Two panels of paired change with 95% confidence intervals for glucocorticoid-responsive signatures in rheumatoid arthritis before and after methotrexate and in lupus nephritis across three time points. All intervals lie below the dashed minimum detectable effect line.

### Boundary conditions: prior specificity and exposure intensity (Supplementary Fig. S3)

In the single-dose prednisolone challenge (GSE111789; purified eosinophils; n=3), only the curated Gene Ontology set separated from the expression-matched null (q=0.12); in the multi-day target-tissue dexamethasone experiment (GSE17992; muscle; n=10), the same set detected exposure as a within-subject specificity contrast (dz=1.24; p=0.006; q=0.018; floor 1.0). Prior specificity and exposure intensity, not dose, governed detectability.

### Positive control: detectability tracks interval, not dose (Fig. 3)

In the sci-Plex 2 series, pseudobulk at well level (six wells per dose) detected the drug-matched dexamethasone prior at dose 100 versus vehicle (dz=+3.70; p < 0.001; floor 1.80), the Gene Ontology set a precise zero and the interferon control near null; cell-level scores reproduced the dose ladder (dz 0.79-1.03 across a 1,000-fold dose range). Across paradigms, the 0.5-2 h window stayed below an n=3 floor (MDE 3.26) while the 96-hour target-tissue contrast exceeded its floor.

**Figure 3.**
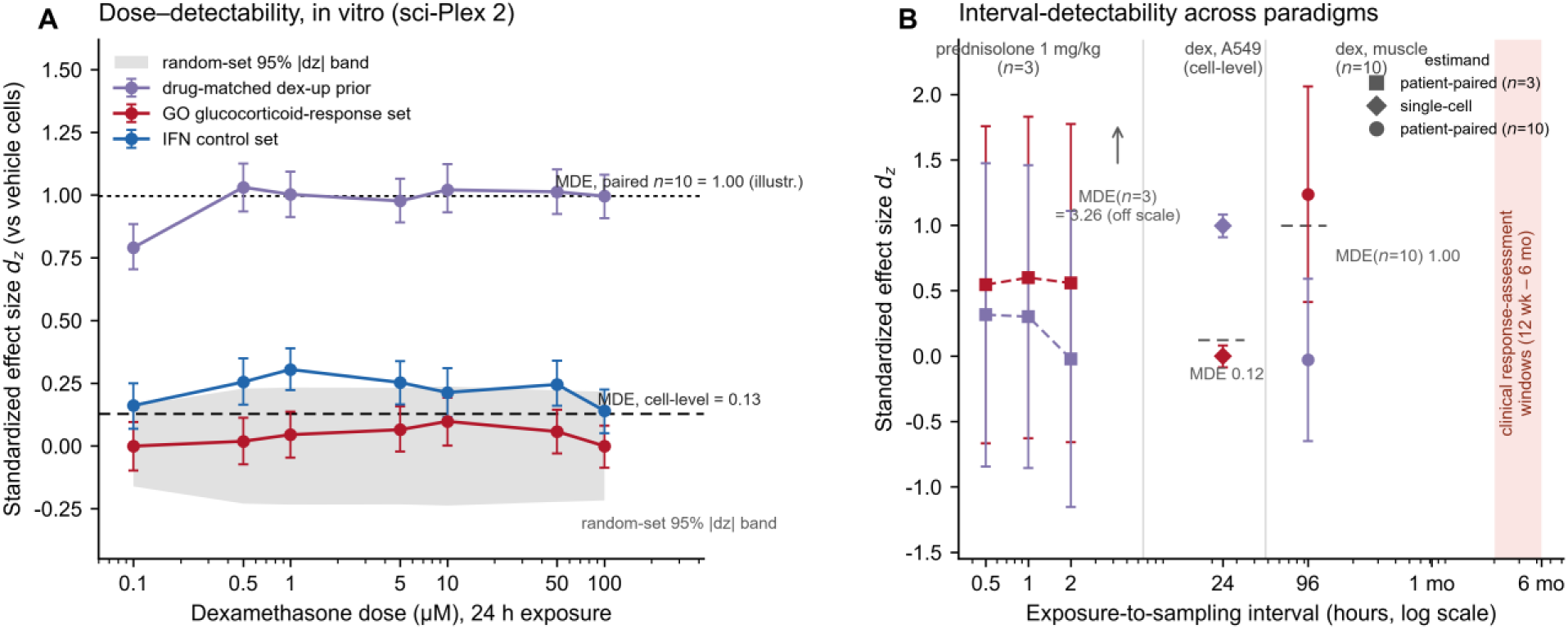
Dose and interval in the positive control. (A) sci-Plex 2 dose series: the well-level pseudobulk contrast is primary (dose 100 vs vehicle, dz = +3.70, floor 1.80); cell-level ladder, shown as sensitivity: drug-matched prior above its floor at every dose (dz = 0.79-1.03), Gene Ontology set null, interferon control near null. (B) Detectability across intervals and paradigms, floors annotated; point shape = estimand (n = 3 / single-cell / n = 10); no extrapolation implied. Alt text: Two panels. A, effect size by dose; the well-level pseudobulk contrast is reported in the text, the drug-matched prior above the cell-level floor. B, effect size by interval across three paradigms, floors marked.

### A benchmark label in current use does not clear its own floor

The ImmunoSTATE trajectory label was highly significant at cell level (p=1.2 × 10-10, pseudo-replicated), but at patient level (dz=-0.97; 95% CI −2.57 to −0.31; 8 of 9 pairs) it did not clear the floor (MDE 1.16); neither drug arm detected its label.

Extended audit of response labels in current use (Table 2, Supplementary Table S4, and Fig. 4) Choi and colleagues’ 39-gene anti-TNF signature [22] was heterogeneous across its eight cohorts: three were nominally significant but did not clear their floors (dz 0.76-1.32; floors 0.90-1.47). Its largest training cohort pointed opposite to the published claim (dz=-0.45; 95% CI −0.82 to −0.07; p=0.016); because |dz|=0.45 lies below that cohort’s own floor of 0.52, the departure is not resolved at the design’s resolution and we score it inconclusive rather than reversed. The pooled estimate was +0.26 (95% CI −0.18 to 0.70; I²=69%) against a pooled floor of 0.31 (Table 2). Widened to a cross-disease panel (Table 2), the IBD comparator was detected as published (dz=-1.51; p=0.006) and one ulcerative colitis cohort, scored with a repurposed diagnostic signature, gave dz=+1.29 (p=0.008) and is reported as a comparator without a verdict. At cohort level, only 1 of 15 auditable cohorts was detected as published; of the other 14, 11 were unresolved at their own floor and 3 bounded below it (Table S5). Three point opposite to the claim, each below its own floor, so no cohort is scored as sign-reversed. That composition is robust to the interval estimator: under percentile, BCa and t-based intervals the detected count is 1 and the reversed count 0 (Table S6.5). Audit-measured effect size ordered the AUCs (|dz| 0.07-1.51 against AUC 0.75-0.96), each exceeding the ceiling its deposited effect can support (Φ(|dz|/√2); Fig. 4, Table S7); as multivariate models can exceed that ceiling, the excess bounds what the label cannot explain. The verdicts do not rest on our reconstruction: of the three cohorts whose deposited split matches the published numbers, none cleared its floor (0 of 3; Table S6). Injection calibration (Fig. S6) gave a 2% null false-positive rate and 54% power at the floor, so below-floor verdicts are conservative.

**Figure 4.**
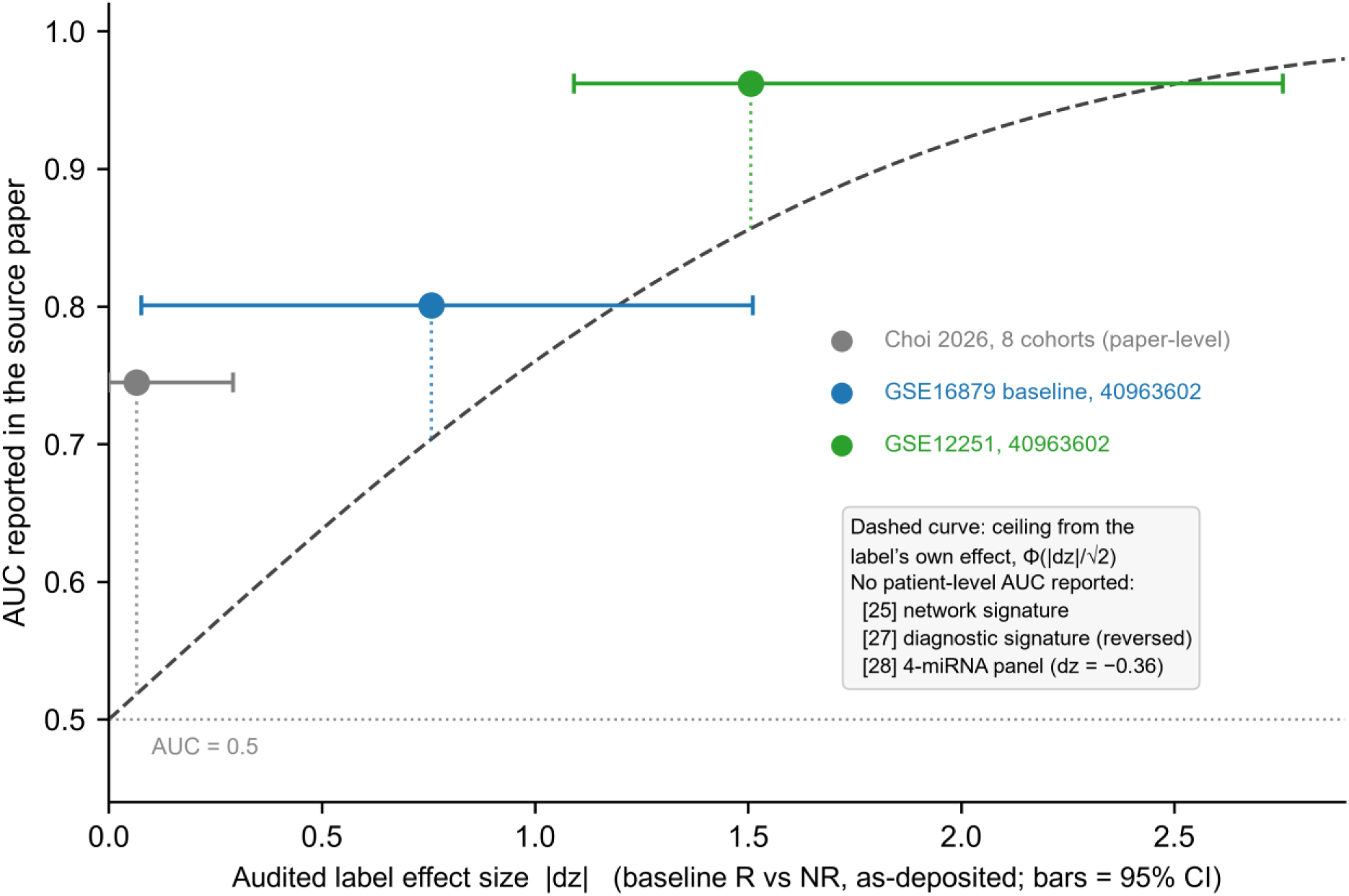
Audit-measured label effect size against reported model performance. Absolute dz (bars, bootstrap 95% CIs) versus the AUC each source paper reported. Dashed curve: ceiling from the label’s own effect, Φ(|dz|/√2); dotted segments, the excess. The first cohort’s CI crosses zero. Cases without a patient-level AUC are in the inset. Alt text: Scatter plot of audited absolute effect size, with bootstrap confidence intervals, against the AUC reported by each source paper. A dashed curve is the ceiling from the label’s own effect and dotted segments show the excess; all three points lie above it. Cohorts without a reported AUC are in an inset.

**Table 2.** Extended audit: verdicts by paper and cohort.

| Paper (disease, label type) | Audited cohorts | dz (key cohort) | Reported AUC | Paper-level verdict |
|---|---|---|---|---|
| [22] RA — 39-gene anti-TNF signature (machine learning) | 8 (5 with a provenance gap flagged in Table S1) | −0.45 (GSE78068, largest); 3 nominally significant at dz 0.76–1.32 | 0.75 (pooled; Table S7) | **F1b**: no cohort detected; pooled +0.26 (−0.18 to 0.70), I² = 69%, against a pooled floor of 0.31 |
| [25] RA — 8-component network signature (1,351 genes) | 6 analysable + 1 F3 (GSE19821) | −0.41 pooled; GSE42296 −1.22 (p = 0.046) | none reported | **F1b (pooled)**: interval spans zero and its pooled floor (0.49) |
| [26] UC — NET 2-gene model (FCGR3B+IL1RN) | GSE12251; GSE16879 (strict baseline) | −1.51 (GSE12251, p = 0.006, as claimed) | 0.96 / 0.80 | **detected** in GSE12251; GSE16879 significant but below floor; pseudoreplication registered |
| [27] UC — diagnostic 4-gene signature repurposed for response | GSE16879 (same cohort as [26]) | +1.29 (p = 0.008) | diagnostic only (0.86–0.97) | **comparator, no verdict**: a diagnostic signature carries no direction expectation for a response task; 28/33 grouping F3 |
| [28] RA — triptolide 4-miRNA panel | GSE106894 | −0.36 (p = 0.55; MDE ≈ 1.65) | none reported | **F1a (bounded below floor)** |
| GSE3592 ([25] test set) | 2006 cDNA array, 8% signature coverage | +0.65 (p = 0.29, reference only) | — | **not assessable** (annotation generation gap) |

Two features bound how far that composition travels. The single detection was the inflammatory bowel disease comparator cohort GSE12251, scored with a two-gene neutrophil-extracellular-trap model (FCGR3B and IL1RN) developed for anti-TNF non-response in that disease: a label matched to the population it was built for does clear its floor, and the detection is informative in that positive direction only, since among the 14 rheumatoid arthritis cohorts the opposite holds. Because 14 of the 15 cohorts derive from two papers, and five of the first paper’s eight cohorts do not permit exact reconstruction of the published subsets, the counts are descriptive rather than binomial; bounding the 30 unaudited records gives 31%-98% for the undetected fraction under any assignment of those records (Fig. S5; margins in Fig. S4).

The pooled estimate is not a stable property of the label either. Recomputing it with a single cohort removed moves it between +0.15 and +0.39, and two exclusions carry the point estimate above its own recomputed floor (Supplementary Tables S3, S5 and S6); both intervals still span zero, so neither exclusion changes any cohort’s verdict. The pooled value sitting below its floor is therefore not independent of whether that cohort is included, and the pooled-below-floor reading should be held with that boundary attached.

Under all scorers the disease-state label was detected and no exposure label reached significance (Fig. S1).

## Discussion

Two findings summarise the results. Within the same patients, tissues and scoring framework, disease-state labels were detectable and treatment-exposure labels were not, in all three clinical cohorts and under every scorer; exposure was measurable only where a specific prior was matched to its cellular context at high intensity. Of the 15 audited response cohorts, one was detectable in the published direction, three bounded below their floors and eleven unresolved at them; three pointed the other way, each below its own floor, none reversed (Supplementary Methods 1.4). What is tested is a label’s detectability **as it is deployed**—as a proxy for the exposure or response state its authors claim it encodes—not its predictive value under another construction: a label with no measurable group difference in a design may still carry multivariate signal this audit does not test. A verdict is a statement about a design, not an allegation about a study; our own positive control was detected only once the design was matched to the effect.

### Drug-specific fingerprints and disease-activity change

One objection: if the interferon signature falls in ten of ten patients after immunosuppression, the design has measured an effect of treatment—any label encoding the state a drug acts on would be expected to move. What such a design does not resolve is the drug-specific component distinguishing glucocorticoid, methotrexate and anti-TNF—small relative to the design’s floor and confounded with baseline activity. Because remission often leaves subclinical inflammation, improvement in a disease-activity label would not in itself indicate that the tissue carried a drug-specific fingerprint.

### Negative results, quantitatively translated

Each null, read against its floor, is a bound: certifying the largest cohort’s magnitude (|dz|=0.45) would require approximately 79 patients per group and the pooled estimate (dz=0.26) approximately 230. With I²=69%, between-cohort heterogeneity rather than sample size appears to be the limiting factor, which would favour design modification over cohort expansion as the next step.

### Relation to sample-size estimation

The floor is not a new statistic—|dz| ≥ MDE is algebraically equivalent to |t| ≈ 2.80 (Supplementary Methods), and that a design too small to detect an effect cannot license a claim about it is long established in the methodological literature [38]. The distinction is one of object: sample-size estimation asks whether a study can detect the effect it intends; this analysis asks whether a label is detectable in the design that defined it, with the margin |dz|/MDE reported as a property of the label.

### Where the spectrum places these results

Conventional response sits where the spectrum predicts: the floor falls with √n while the displacement is fixed by biology, so design features rather than cohort size would move a label along it. Anifrolumab, which blocks the pathway its own label measures, is one clinical instance at the other end [39].

### Weak labels amplify method choices into artefacts

Z-score-mean scoring tracks a sample’s global expression level, inflating signature correlations in mixed designs (Fig. S2); under rank-based scoring, a post-methotrexate TNF increase (empirical p=0.005) did not reproduce in the authors’ own analysis (p=0.73), consistent with a scoring artefact. Where the underlying effect is small, such artefacts may determine the interpretation. Direction is therefore read conservatively: where a cohort points opposite to the published claim, that departure is below the cohort’s floor, and baseline-state confounding is one reading rather than a demonstrated mechanism (Supplementary Methods 1.9).

### What the margin means for a label in use

The practical implication is a reporting proposal rather than a new statistic: before a response label is used to stratify patients or benchmark a model, two quantities could be reported—the label’s own effect size in the design that produced it, and the minimum detectable effect of that design. Their ratio, |dz|/MDE, is the fraction of the design’s resolving power that the observed effect occupies; below 1, the design would not have been expected to detect the label at that magnitude. For the labels audited here that ratio is 0.84 at the pooled level, below 1 for all 14 audited rheumatoid arthritis cohorts (range 0.03-0.90) and 1.23 for the one comparator that cleared its floor. This comparison is available before any data are collected: a trialist can compare a planned design’s minimum detectable effect with the effect size of the label they intend to use, and a reader can apply the same comparison to any published label.

### Implications for reporting and design

Three changes follow from these results, offered as proposals: reporting the design floor alongside any null; reporting a response label’s effect size before any benchmark that uses it; and treating a below-floor label as untested for stratification purposes, since a model’s reported performance cannot by itself establish that the label was measurable. The proposed reporting set is given in the Supplementary Methods (label detectability reporting checklist); Tables S8 and S9 give the floor for each design here and three published nulls read against theirs.

### Limitations

Sample sizes are small (10-28 pairs) but are the paired cohorts that exist. Bulk data lack single-cell resolution, so cell-confined effects may be diluted below the floor [40]. Responder status rests on change quantities and baseline activity is partly collinear with change, so direction is conservative. Irreproducible labels are scored F3, excluded and not attributed to any analysis. The 14 non-detections rest on two source papers, and intervals are percentile bootstrap, whose coverage at the smallest split was 89% against a nominal 95%: that shortfall is quantified, and verdicts were recomputed under BCa and t-based intervals, the detected count remaining 1 and the reversed count 0 (Table S6.5). The IBD layer does not license extrapolation, and detectability is relative to the priors and scorers used.

## Conclusions

In the cohorts examined here, at conventional treatment intensities and routine sampling intervals, exposure and response-prediction labels did not reach the detection floors of the designs that define them, whereas disease-state labels in the same samples did. Designs with target tissue, purified cells or mechanism-specific exposure did move labels above their floors. Reporting both a response label’s effect size and the minimum detectable effect of its design would allow the comparison before the label is used for stratification or benchmarking.

## Data Availability

All datasets analysed are publicly available; accessions and designs are listed in Table 1 and
annotation provenance for the extended audit in Supplementary Table S1. Analysis scripts,
adjudication ledgers, per-sample mapping tables and the pre-registered verdict taxonomy are
deposited at Zenodo (doi:10.5281/zenodo.23123981), with restricted access and a private review
link for reviewers. The pre-registration predates every verdict reported.

## Declarations

## Ethics approval and consent to participate

This study re-analysed publicly available, de-identified transcriptomic data from public repositories; no new human or animal subjects were recruited and no identifiable personal data were handled. The original studies were conducted under the ethics approvals reported in their respective publications. Ethical approval was therefore not required for this re-analysis, because it involved no recruitment, no intervention and no access to identifiable personal data, and consent to participate is likewise not applicable: all data were obtained from public repositories in de-identified form, and no individual patient data, images or identifiers are reported.

## Consent for publication

Not applicable: the manuscript and its figures contain no identifiable patient material.

## Availability of data and materials

All datasets analysed are publicly available and are cited in the reference list [41–48]; accessions, annotation provenance, download dates and per-cohort label-mapping rules are listed in Supplementary Table S1 and the deposited code package. Analysis scripts, adjudication ledgers, per-sample mapping tables, the pre-registered verdict taxonomy and all result files are deposited at Zenodo (doi:10.5281/zenodo.23123981), with restricted access and a private review link supplied for reviewers; the pre-registration predates every verdict reported. No new data were generated; all input data came from public repositories under their licences, and no controlled-access material was used. The underlying numerical results for every display item are supplied in the supplementary tables and in the deposited package, so each figure and table can be reconstructed from public data without further access.

## Authors’ contributions

Conceptualization: Haitao Yang, Yan Huang. Methodology: Haitao Yang, Yan Huang. Formal analysis: Haitao Yang, Yan Huang. Investigation: Jianghua Xu, Suo Zhang, Rui Liao. Data curation: Yan Huang. Writing – original draft: Haitao Yang, Yan Huang. Writing – review and editing: all authors. Visualization: Jianghua Xu, Rui Liao. Supervision: Haitao Yang. Project administration: Haitao Yang. All authors read and approved the final manuscript.

## AI Disclosure Statement

This manuscript was prepared with the assistance of large language model-based writing assistants: DeepSeek V4 and Kimi K3. These tools were used for English-language editing, for compression to the journal’s word limit, and for assistance with analysis code. They were not used to generate data, results, figures, tables or references. Every number reported here was computed by the deposited code from the datasets listed in Table 1 and verified by the authors against those outputs. All content generated or modified with these tools was checked and approved by the authors, who accept full responsibility for its accuracy, integrity and originality. The tools are not authors.

## References

1. Bennett L, Palucka AK, Arce E, et al. Interferon and granulopoiesis signatures in systemic lupus erythematosus blood. J Exp Med. 2003;197(6):711–723. doi:10.1084/jem.20021553. PMID: 12642603.

2. Taroni JN, Martyanov V, Mahoney JM, Whitfield ML. A functional genomic meta-analysis of clinical trials in systemic sclerosis: toward precision medicine and combination therapy. J Invest Dermatol. 2017;137(5):1033–1041. PMID: 28011145.

3. Northcott M, Jones S, Koelmeyer R, et al. Type 1 interferon status in systemic lupus erythematosus: a longitudinal analysis. Lupus Sci Med. 2022;9(1):e000625. doi:10.1136/lupus-2021-000625. PMID: 35197305.

4. Dolcino M, Tinazzi E, Pelosi A, et al. Gene expression analysis before and after treatment with adalimumab in patients with ankylosing spondylitis identifies molecular pathways associated with response to therapy. Genes (Basel). 2017;8(4):127. doi:10.3390/genes8040127. PMID: 28441778.

5. Moysidou GS, Garantziotis P, Sentis G, et al. Molecular basis for the disease-modifying effects of belimumab in systemic lupus erythematosus and molecular predictors of early response. Ann Rheum Dis. 2025;84(2):262–273. doi:10.1136/ard-2024-226051. PMID: 39919899.

6. Cui H, Wang C, Maan H, et al. scGPT: toward building a foundation model for single-cell multi-omics using generative AI. Nat Methods. 2024;21(8):1470–1480. doi:10.1038/s41592-024-02201-0.

7. Theodoris CV, Xiao L, Chopra A, et al. Transfer learning enables predictions in network biology. Nature. 2023;618:616–624. doi:10.1038/s41586-023-06139-9.

8. Hao M, Gong J, Zeng X, et al. Large-scale foundation model on single-cell transcriptomics. Nat Methods. 2024;21(8):1481–1491. doi:10.1038/s41592-024-02305-7.

9. Sun R, Qian L, Li Y, et al. An operational perturbation proteomics-based virtual cell model. Nature. 2026. doi:10.1038/s41586-026-11001-9. PMID: 42717098. Published online 2026 Sep 9.

10. Ahlmann-Eltze C, Huber W, Anders S. Deep-learning-based gene perturbation effect prediction does not yet outperform simple linear baselines. Nat Methods. 2025;22(8):1657–1661. doi:10.1038/s41592-025-02772-6. PMID: 40759747.

11. Wilkinson MGL, Moulding D, McDonnell TCR, et al. Role of CD14+ monocyte-derived oxidised mitochondrial DNA in the inflammatory interferon type 1 signature in juvenile dermatomyositis. Ann Rheum Dis. 2023;82(5):658–669. doi:10.1136/ard-2022-223469. PMID: 36564154.

12. Greenberg SA, Sanoudou D, Haslett JN, et al. Molecular profiles of inflammatory myopathies. Ann Neurol. 2005;57(5):664–678. doi:10.1002/ana.20487.

13. Chen YW, Shi R, Geraci N, et al. Duration of chronic inflammation alters gene expression in muscle from untreated girls with juvenile dermatomyositis. BMC Immunol. 2008;9:43. doi:10.1186/1471-2172-9-43.

14. Gene Expression Omnibus. GSE193276: paired skin biopsies from adult dermatomyositis patients before and after apremilast add-on therapy. https://www.ncbi.nlm.nih.gov/geo/query/acc.cgi?acc=GSE193276 (accessed 2026-09-23).

15. Shoda J, Etori K, Hattori K, et al. Semaphorin 3G exacerbates joint inflammation through the accumulation and proliferation of macrophages in the synovium. Arthritis Res Ther. 2022;24(1):134. doi:10.1186/s13075-022-02817-7. PMID: 35659346.

16. Ducreux J, Houssiau FA, Vandepapelière P, et al. Interferon α kinoid induces neutralizing anti-interferon α antibodies that decrease the expression of interferon-induced and B cell activation associated transcripts: analysis of extended follow-up data from the interferon α kinoid phase I/II study. Rheumatology (Oxford). 2016;55(10):1901–1905. doi:10.1093/rheumatology/kew262. PMID: 27354683.

17. Khoury P, Liter O, Cahill KN, et al. Single-dose prednisone effect on gene expression in blood eosinophils. Allergy. 2018;73(10):2076–2079. doi:10.1111/all.13497.

18. Gadkari M, Coetzer M, Papazoglou A, et al. A functional genomic analysis of corticosteroid response in eosinophils from patients with hypereosinophilic syndromes. Sci Data. 2018;5:180275. doi:10.1038/sdata.2018.275.

19. Viguerie N, Picard F, Hul G, et al. Gene expression analysis in human skeletal muscle and adipose tissue following short-term glucocorticoid treatment. Physiol Genomics. 2012;44(2):141–151. doi:10.1152/physiolgenomics.00032.2011.

20. Srivatsan SR, McFaline-Figueroa JL, Ramani V, et al. Massively multiplex chemical transcriptomics at single-cell resolution. Science. 2020;367(6473):45–51. doi:10.1126/science.aax6234. PMID: 31806696.

21. Peidli S, Green TD, Shen C, et al. scPerturb: harmonized single-cell perturbation data. Nat Methods. 2024;21(3):531–540. doi:10.1038/s41592-023-02144-y. PMID: 38279009.

22. Choi Y, Kim T, Kim HJ, Lee D, Lee KE. Blood-based gene signatures associated with therapeutic response to anti-TNF therapy in rheumatoid arthritis: a combined meta-analytical and machine learning approach. EULAR Rheumatol Open. 2026;2(1). doi:10.1016/j.ero.2025.11.022. PMID: 42370093.

23. Toro-Domínguez D, Martorell-Marugán J, Martinez-Bueno M, et al. Scoring personalized molecular portraits identify systemic lupus erythematosus subtypes and predict individualized drug responses, repositioning and drug synergy. Briefings in Bioinformatics. 2022;23(5). doi:10.1093/bib/bbac332. PMID: 35947992.

24. Lee DJ, Tsai PH, Chen CC, Dai YH. Incorporating knowledge of disease-defining hub genes and regulatory network into a machine learning-based model for predicting treatment response in lupus nephritis after the first renal flare. J Transl Med. 2023;21(1). doi:10.1186/s12967-023-03931-z. PMID: 36737814.

25. Thomson TM, Lescarbeau RM, Drubin DA, et al. Blood-based identification of non-responders to anti-TNF therapy in rheumatoid arthritis. BMC Med Genomics. 2015;8:34. doi:10.1186/s12920-015-0100-6. PMID: 26036272.

26. Ou J, Li L, Zhi F, Huang B, Zhao X. Developing models for the diagnosing of ulcerative colitis and prognosis of anti-TNF-α non-response based on neutrophil extracellular trap-associated genes. Front Immunol. 2025;16:1530508. doi:10.3389/fimmu.2025.1530508. PMID: 40963602.

27. Wang M, Liang L, Tang Z, Han J, Wu L, Liu L, Chen Y. Multi-omics derivation of a core gene signature for predicting therapeutic response and characterizing immune dysregulation in inflammatory bowel disease. Front Immunol. 2025;16:1611598. doi:10.3389/fimmu.2025.1611598. PMID: 40821793.

28. Zhang Y, Wang H, Mao X, et al. A novel circulating miRNA-based model predicts the response to Tripterygium Glycosides Tablets: moving toward model-based precision medicine in rheumatoid arthritis. Front Pharmacol. 2018;9:378. doi:10.3389/fphar.2018.00378. PMID: 29881347.

29. Mendoza-Pinto C, Sánchez-Tecuatl M, Berra-Romani R, et al. Machine learning in the prediction of treatment response in rheumatoid arthritis: a systematic review. Semin Arthritis Rheum. 2024;68:152501. doi:10.1016/j.semarthrit.2024.152501. PMID: 39226650.

30. Gehringer CK, Martin GP, Hyrich KL, et al. Clinical prediction models for methotrexate treatment outcomes in patients with rheumatoid arthritis: a systematic review and meta-analysis. Semin Arthritis Rheum. 2022;56:152076. doi:10.1016/j.semarthrit.2022.152076. PMID: 35921745.

31. Liberzon A, Birger C, Thorvaldsdóttir H, et al. The Molecular Signatures Database (MSigDB) hallmark gene set collection. Cell Syst. 2015;1(6):417–425. doi:10.1016/j.cels.2015.12.004. PMID: 26771021.

32. Ashburner M, Ball CA, Blake JA, et al. Gene ontology: tool for the unification of biology. Nat Genet. 2000;25(1):25–29. doi:10.1038/75556. PMID: 10802651.

33. Kuleshov MV, Jones MR, Rouillard AD, et al. Enrichr: a comprehensive gene set enrichment analysis web server 2016 update. Nucleic Acids Res. 2016;44(W1):W90–W97. doi:10.1093/nar/gkw377. PMID: 27141961.

34. Moncrieffe H, Hinks A, Ursu S, et al. Generation of novel pharmacogenomic candidates in response to methotrexate in juvenile idiopathic arthritis: correlation between gene expression and genotype. Pharmacogenet Genomics. 2010;20(11):665–676. doi:10.1097/FPC.0b013e32833f2cd0. PMID: 20827233.

35. Foroutan M, Bhuva DD, Lyu R, et al. Single sample scoring of molecular phenotypes. BMC Bioinformatics. 2018;19(1):404. doi:10.1186/s12859-018-2435-4. PMID: 30400809.

36. Nakanishi K, Shimizu H. Integrating virtual cell modeling with patient-derived transcriptomics to uncover target signals in rheumatoid arthritis. bioRxiv. 2026. doi:10.64898/2026.04.19.719532. Preprint (not peer-reviewed).

37. He C, Ren Z, Xia X, et al. A single-cell RNA-seq dataset of synovial fluid from rheumatoid arthritis treated with TNF-α/JAK inhibitor. Sci Data. 2025;12(1):977. doi:10.1038/s41597-025-05339-4. PMID: 40494845.

38. Button KS, Ioannidis JP, Mokrysz C, et al. Power failure: why small sample size undermines the reliability of neuroscience. Nat Rev Neurosci. 2013;14(5):365–376. doi:10.1038/nrn3475. PMID: 23571845.

39. Morand EF, Furie R, Tanaka Y, et al. Trial of anifrolumab in active systemic lupus erythematosus. N Engl J Med. 2020;382:170–180. doi:10.1056/NEJMoa1912196. PMID: 31851795.

40. Song W, Lee EE, Park S, et al.; Single-Cell Atlas of Immune Diseases Consortium. Type 1 interferon signature and allograft inflammatory factor-1 contribute to refractoriness to TNF inhibition in ankylosing spondylitis. Nat Commun. 2025;16(1):5531. doi:10.1038/s41467-025-60445-6. PMID: 40593526.

41. Gene Expression Omnibus: GSE221091. https://www.ncbi.nlm.nih.gov/geo/query/acc.cgi?acc=GSE221091

42. Gene Expression Omnibus: GSE176440. https://www.ncbi.nlm.nih.gov/geo/query/acc.cgi?acc=GSE176440

43. Gene Expression Omnibus: GSE72747. https://www.ncbi.nlm.nih.gov/geo/query/acc.cgi?acc=GSE72747

44. Gene Expression Omnibus: GSE111789. https://www.ncbi.nlm.nih.gov/geo/query/acc.cgi?acc=GSE111789

45. Gene Expression Omnibus: GSE17992. https://www.ncbi.nlm.nih.gov/geo/query/acc.cgi?acc=GSE17992

46. Gene Expression Omnibus: GSE1551. https://www.ncbi.nlm.nih.gov/geo/query/acc.cgi?acc=GSE1551

47. Gene Expression Omnibus: GSE11971. https://www.ncbi.nlm.nih.gov/geo/query/acc.cgi?acc=GSE11971

48. Gene Expression Omnibus: GSE193276. https://www.ncbi.nlm.nih.gov/geo/query/acc.cgi?acc=GSE193276

