## Supplementary Figures S1-S6 for "A detectability floor for treatment-response labels in transcriptomics: a methodological audit using rheumatoid arthritis as a test case"

*Supplementary material to: "A detectability floor for treatment-response labels in transcriptomics: a methodological audit using rheumatoid arthritis as a test case"*

**Figure S1**

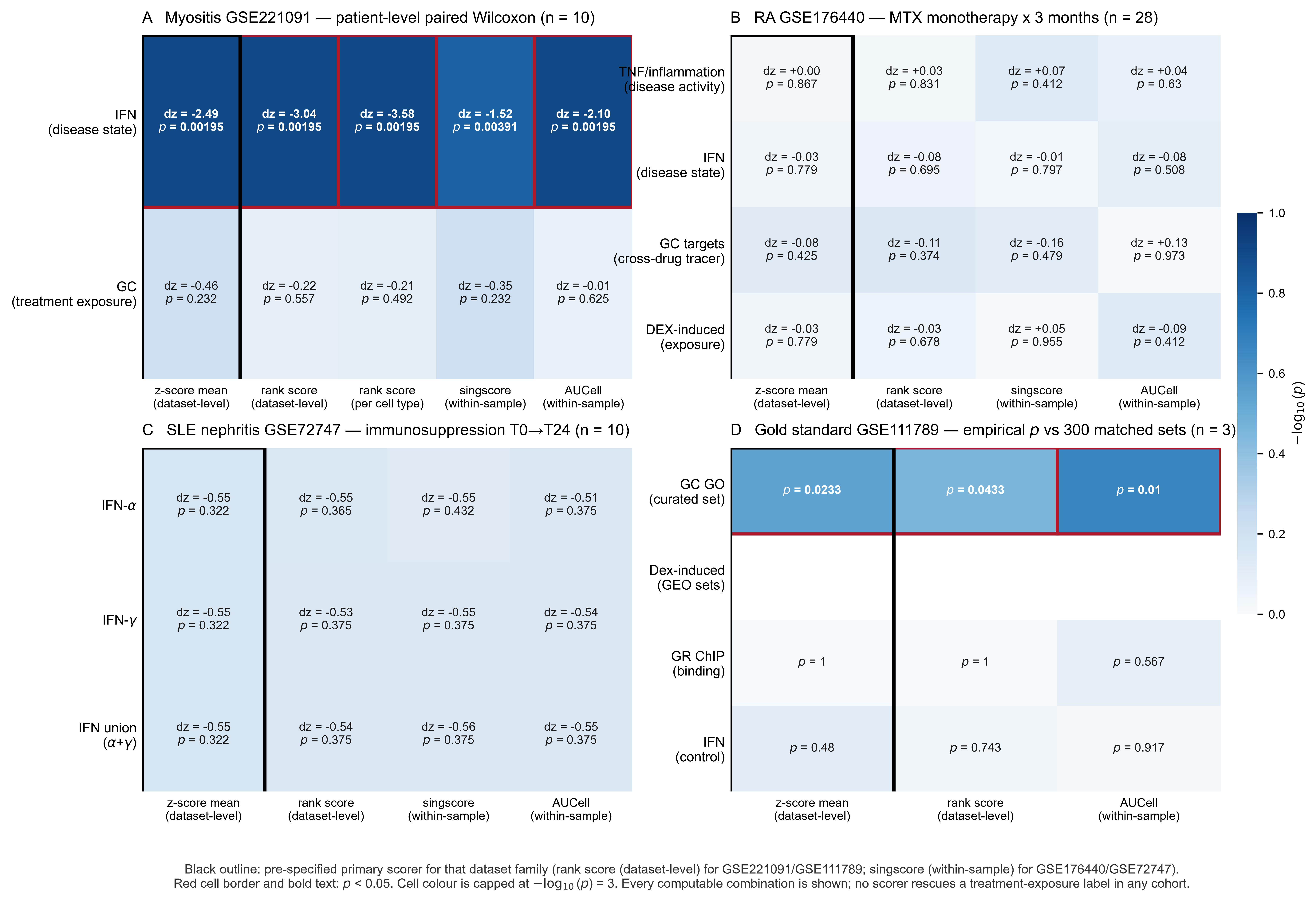

**Figure S1. Scorer robustness.** Every cohort x signature combination under all five scorers: the disease-state label is detected under every scorer and no exposure label reaches significance under any.

Alt text: Heat map of results for every cohort and signature combination under five scoring methods, showing detection of the disease-state label under all methods and no significant exposure label under any method.

**Figure S2**

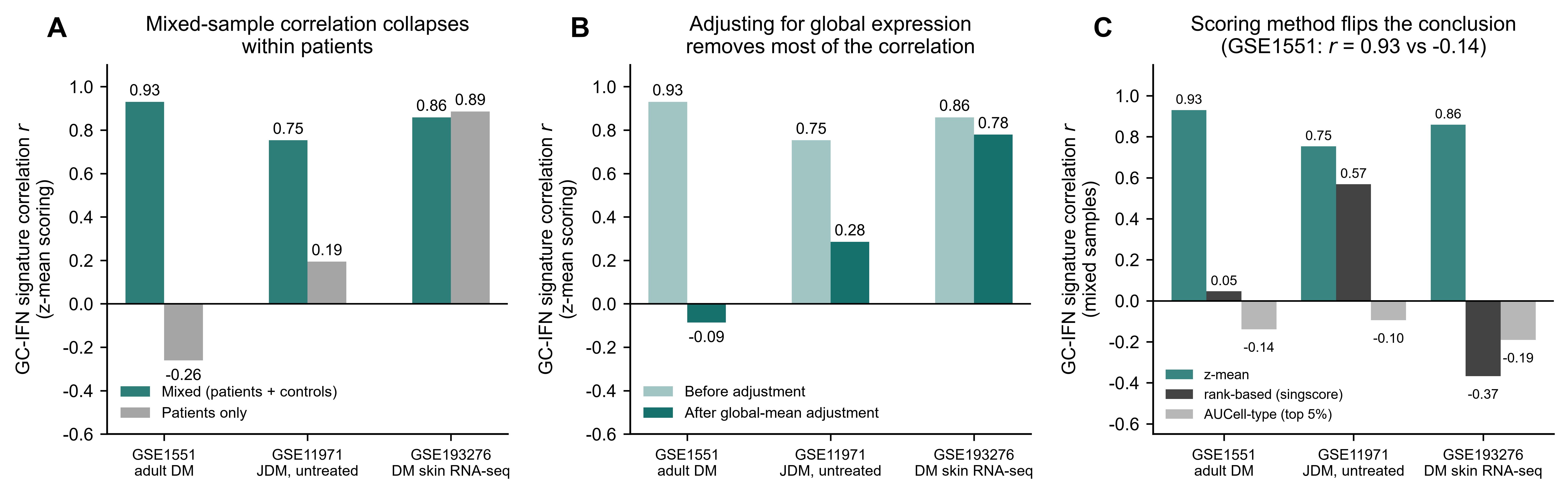

**Figure S2. Mixed-sample artefact.** Glucocorticoid-interferon correlations in mixed and within-patient scopes under three scorers, before and after regressing out each sample's global expression level.

Alt text: Correlation coefficients between glucocorticoid and interferon scores under three scoring methods, in mixed samples and within patients, before and after adjustment for each sample's global expression level; strong mixed-sample correlations collapse within patients.

**Figure S3**

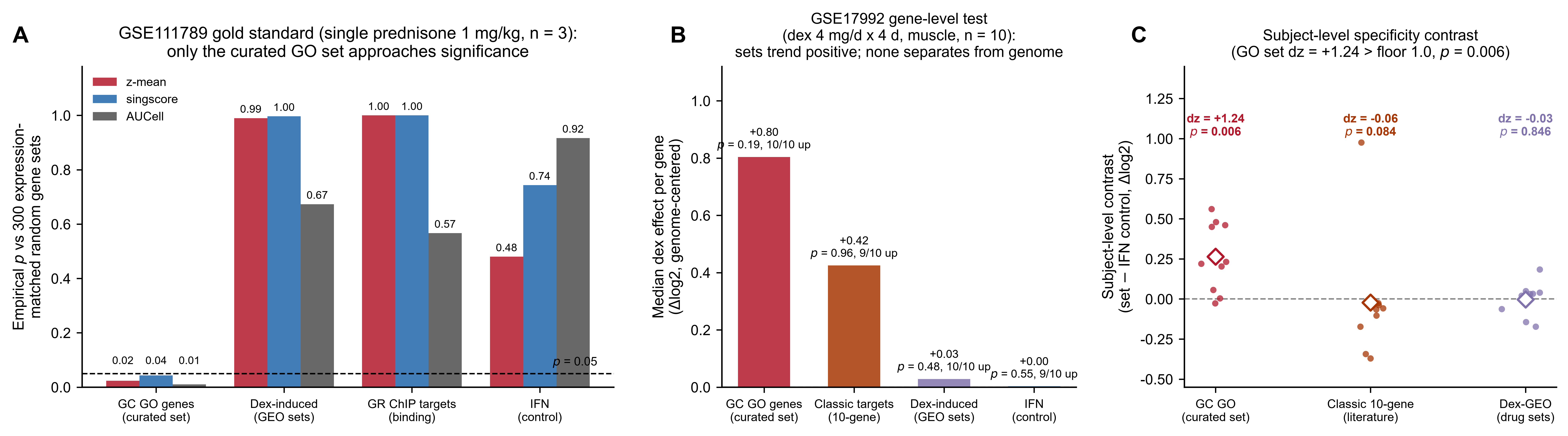

**Figure S3. Boundary conditions.** Empirical separation of curated and less-curated prior sets from expression-matched nulls in the single-dose experiment, and detection of the curated set as a within-subject specificity contrast in the multi-day target-tissue experiment.

Alt text: Two panels. Left, empirical p values for curated and less-curated gene sets against expression-matched nulls in the single-dose prednisolone experiment; only the curated set separates. Right, paired within-subject change for the curated set in the multi-day target-tissue dexamethasone experiment, exceeding its detection floor.

**Figure S4**

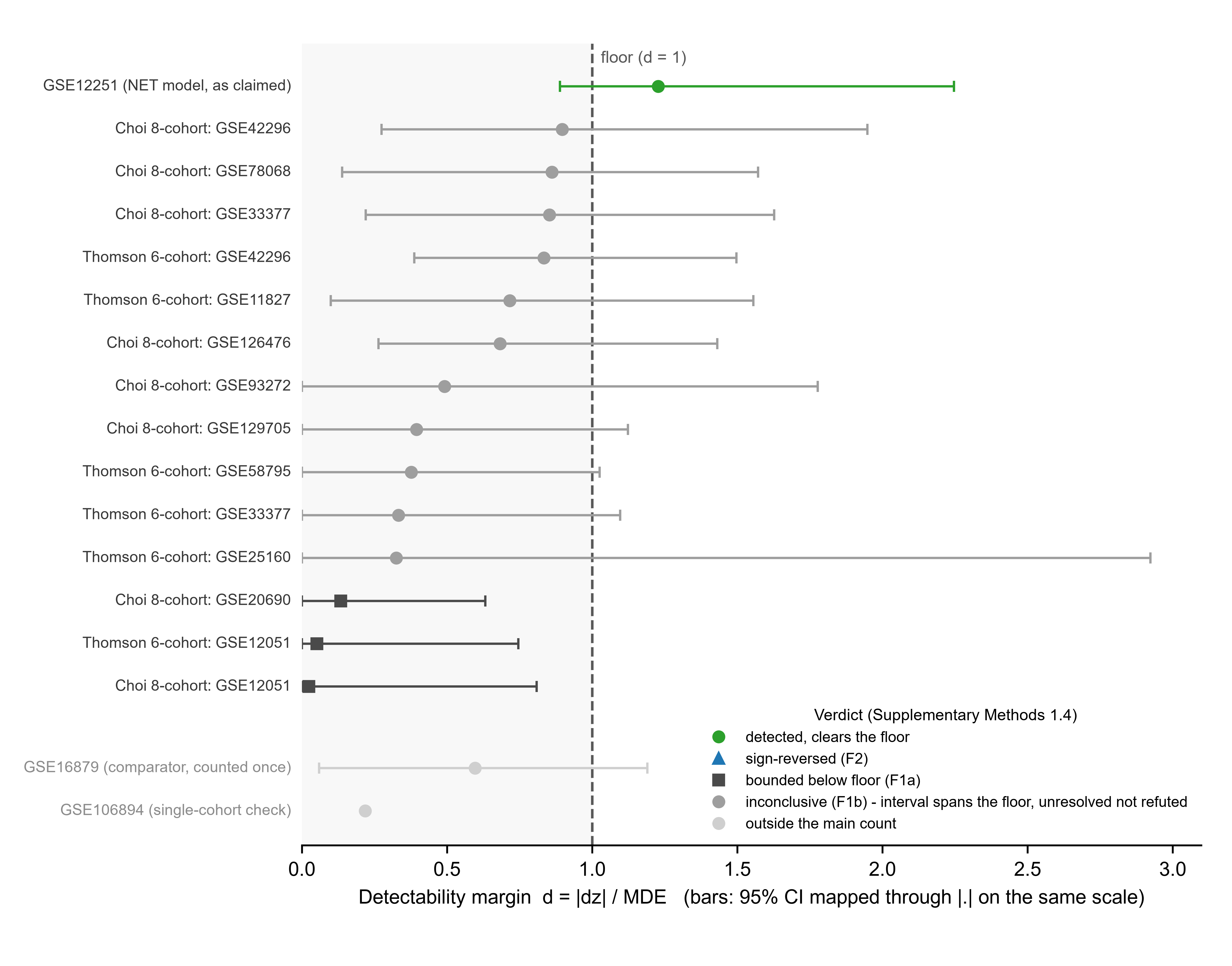

**Figure S4. Detectability margins across the audit.** Detectability margin d = |dz| / MDE per cohort in the main count (Choi eight, Thomson six, GSE12251) and two excluded; d = 1 is the floor. Three classes (Methods 1.4): detected as published (one, d = 1.23); bounded below floor (three); inconclusive (eleven). Three cohorts point opposite to the published claim, each below its own floor, and count as inconclusive: none is scored as sign-reversed.

Alt text: Forest plot of the detectability margin for seventeen audited cohorts, most below the floor line at d = 1. One is detected; three bounded below the floor and eleven inconclusive.

**Figure S5**

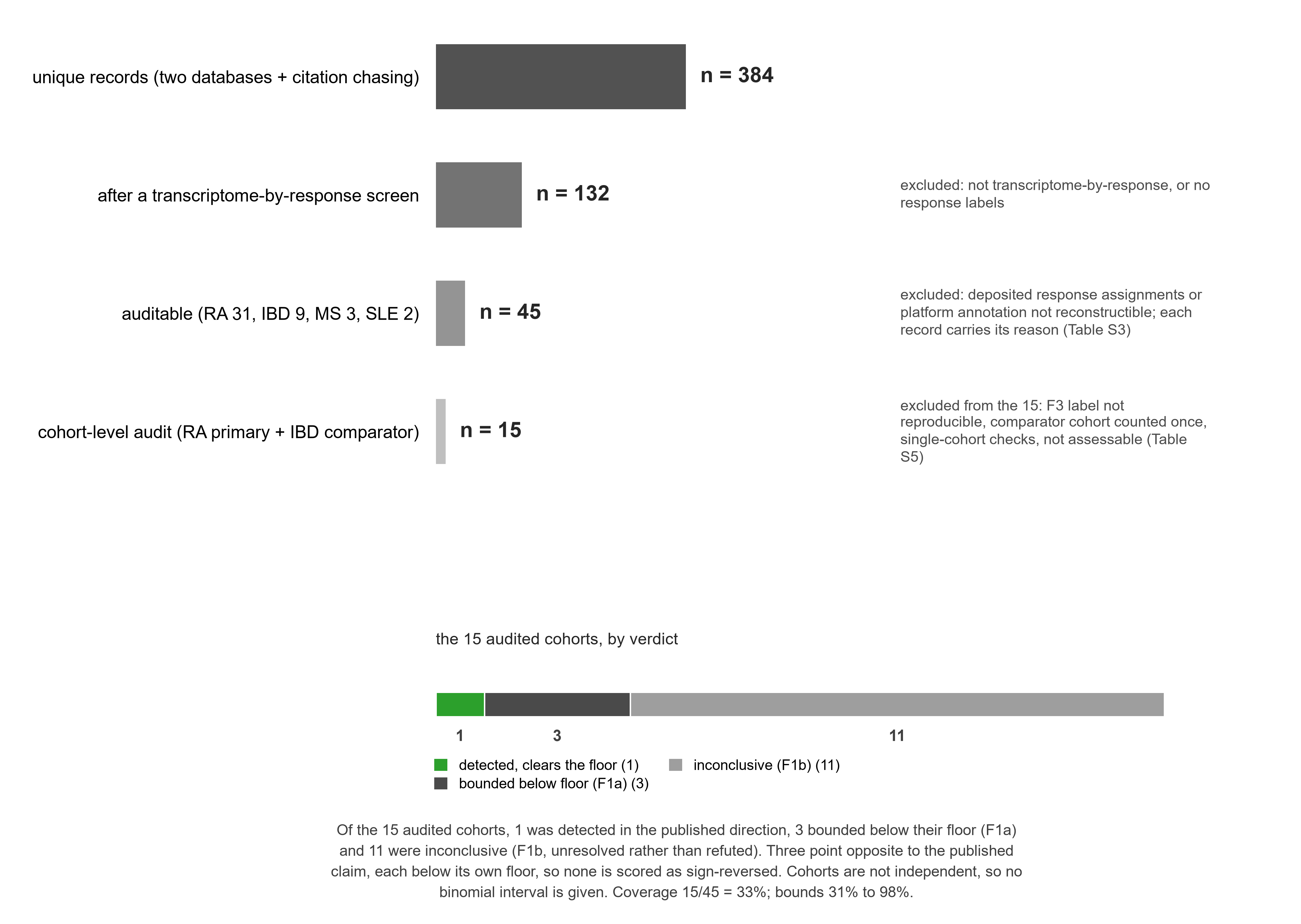

**Figure S5. Complete audit funnel.** From 384 unique records to 132 after a transcriptome-by-response screen, 45 auditable and 15 audited at cohort level. Of the 15, 1 was detected as published, 3 bounded below their floor and 11 unresolved; three point opposite to the claim, each below its own floor, so none is sign-reversed. Coverage 15/45 = 33%; bounding the 30 unaudited gives 31% to 98%. Cohorts are not independent, so no binomial interval (Methods 1.7).

Alt text: Four-step funnel from 384 records to 15 audited cohorts with exclusion reasons, the verdict breakdown, coverage and sensitivity bounds.

**Figure S6**

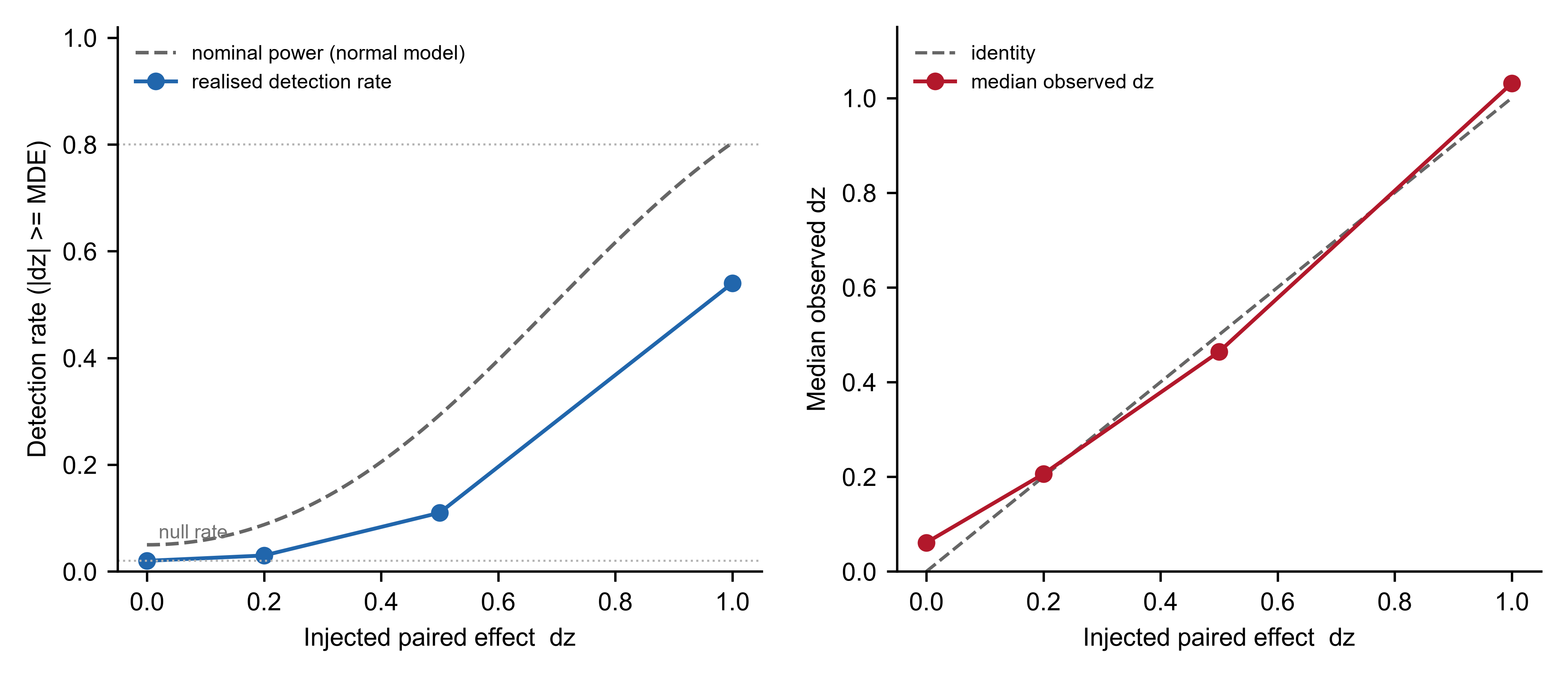

**Figure S6. Injection calibration of the scorer-floor pipeline.** In the real JDM matrix, 100 random 30-gene sets per level were scored and their paired deltas shifted by dz = 0, 0.2, 0.5 or 1.0. Realised detection rates were 2%, 3%, 11% and 54%; the nominal normal-model power at the floor is 80%, the shortfall reflecting heavier-than-normal score distributions.

Alt text: Two panels. Detection rate and median observed effect size against the injected effect, showing a two percent null rate and a realised power of 54 percent at the floor.
